# The Clinical Genome Resource (ClinGen) Familial Hypercholesterolemia Variant Curation Expert Panel consensus guidelines for *LDLR* variant classification

**DOI:** 10.1101/2021.03.17.21252755

**Authors:** Joana R. Chora, Michael A. Iacocca, Lukas Tichy, Hannah Wand, C. Lisa Kurtz, Heather Zimmermann, Annette Leon, Maggie Williams, Steve E. Humphries, Amanda J. Hooper, Mark Trinder, Liam R. Brunham, Alexandre Costa Pereira, Cinthia E. Jannes, Margaret Chen, Jessica Chonis, Jian Wang, Serra Kim, Tami Johnston, Premysl Soucek, Michal Kramarek, Sarah E. Leigh, Alain Carrie, Eric J. Sijbrands, Robert A. Hegele, Tomas Freiberger, Joshua W. Knowles, Mafalda Bourbon On behalf of the ClinGen Familial Hypercholesterolemia Expert Panel

## Abstract

**Purpose:** In 2015, the American College of Medical Genetics and Genomics (ACMG) and the Association for Molecular Pathology (AMP) published consensus standardized guidelines for variant classification in Mendelian disorders. To increase accuracy and consistency, the Clinical Genome Resource (ClinGen) Familial Hypercholesterolemia (FH) Variant Curation Expert Panel (VCEP) was tasked with optimizing the existing ACMG/AMP framework for disease-specific classification in FH. Here, we provide consensus recommendations for the most common FH-causing gene, *LDLR*, where >2,300 unique FH-associated variants have been identified.

**Methods:** The multidisciplinary FH VCEP met in person and through frequent emails and conference calls to develop *LDLR*-specific modifications of ACMG/AMP guidelines. Through iteration, pilot testing, debate and commentary, consensus among experts was reached.

**Results:** The consensus *LDLR* variant modifications to existing ACMG/AMP guidelines include: 1) alteration of population frequency thresholds; 2) delineation of loss-of-function variant types; 3) functional study criteria specifications; 4) co-segregation criteria specifications; and 5) specific use and thresholds for *in silico* prediction tools, among others.

**Conclusion:** Establishment of these guidelines as the new standard in the clinical laboratory setting will result in a more evidence-based, harmonized method for *LDLR* variant classification worldwide, thereby improving the care of FH patients.

## INTRODUCTION

Familial hypercholesterolemia (FH) (OMIM: 143890) is a common (∼1:250 individuals affected)^1^ genetic dyslipidemia characterized by lifelong exposure to elevated low-density lipoprotein cholesterol (LDL-C) levels. Early identification and appropriate treatment are imperative for prevention of premature atherosclerotic cardiovascular disease; however, less than 10% of individuals with FH worldwide have been diagnosed.^2,3^

FH is predominantly caused by heterozygous variants in one of three genes: the LDL receptor gene (*LDLR; >*90% of molecularly defined cases), the apolipoprotein B gene (*APOB; ∼*5-8% of cases), or the proprotein convertase subtilisin/kexin type 9 gene (*PCSK9; ∼*1% of cases).^4^ A single variant in the apolipoprotein E gene (*APOE;* p.Leu167del) is also known to cause the FH phenotype, and recent evidence suggests this variant may explain 1-2% of FH cases in some countries.^5^ Identification of a pathogenic variant in an FH-associated gene can strongly affirm a diagnosis, motivates and simplifies family-based ‘cascade screening’, has potential to direct therapeutic strategy and/or promote adherence, and may impact insurance coverage of certain medications. As genomic technologies – namely next-generation sequencing – have become more affordable and available in recent years, genetic testing has increasingly become a central part of diagnosing FH in many countries. Genetic testing in FH is recommended by the United Kingdom National Institutes for Health and Clinical Excellence,^6^ both the European and International Atherosclerosis Societies,^2^ and an international expert panel convened by the FH Foundation and American College of Cardiology,^7^ among others. The US Centers for Disease Control and Prevention Office of Public Health Genomics also recommends the use of genetic information in the care of FH.^8^ Moreover, the American College of Medical Genetics and Genomics (ACMG) list *LDLR, APOB*, and *PCSK9* among the 59 ‘medically actionable’ genes,^9^ which has, in part, led to frequent inclusion of these genes on commercially available clinical panels,^10^ as well as direct-to-consumer tests.

With the increasingly widespread implementation of genetic testing for FH, it is becoming ever more essential to establish a consensus, standardized method for the clinical classification of identified variants. In a 2018 study of >6,500 FH-associated variants submitted to the ClinVar database, there were at least 12 different variant classification criteria being used among 30 submitters from 14 different countries.^11^ This heterogeneity leads to discordance in variant classification. For instance, 379 unique FH variants had conflicting classifications in ClinVar (i.e., benign/likely benign + uncertain significance; pathogenic/likely pathogenic + uncertain significance; or even benign/likely benign + pathogenic/likely pathogenic).

Application of the American College of Medical Genetics and Genomics / Association for Molecular Pathology (ACMG/AMP) guidelines^12^ has been a major advancement toward achieving a more critical and consistent approach to variant classification for many disorders, including FH.^11^ However, because these guidelines are meant to be generalizable to all Mendelian disorders, they include inherent ambiguities that may lead to differences in their classification and application among users. Indeed, 114 FH-associated variants in ClinVar have conflicting classifications (as defined above) despite each laboratory having cited the same ACMG/AMP guidelines as their applied criteria. Gene-specific modifications to these guidelines are essential to provide the clarity required for standardized variant classification.

In 2013, the Clinical Genome Resource (ClinGen) Consortium was established as a centralized collaborative resource that aims to define the clinical relevance of genes and variants.^13^ Among their major initiatives is the commission of disease/gene expert panels to provide consensus specifications of ACMG/AMP criteria. The ClinGen FH Variant Curation Expert Panel (VCEP) has been tasked with providing gene-specific recommendations for *LDLR, APOB* and *PCSK9*. Here, we describe consensus ACMG/AMP specifications for the *LDLR* gene, where more than 2,300 unique variants have been identified in patients with a clinical association of FH.^11^

## MATERIALS AND METHODS

### ClinGen FH Variant Curation Expert Panel

FH VCEP membership includes clinicians, laboratory diagnosticians, research scientists, genomic medicine specialists, and genetic counsellors, who share expertise knowledge in FH. Members represent a range of academic, clinical and diagnostic laboratory settings. To achieve international harmonization of variant classification practice, additional emphasis was placed on global representation with members coming from 12 countries (United States, Canada, Brazil, United Kingdom, Portugal, Spain, France, Netherlands, Czech Republic, Japan, Australia, and Israel). The FH VCEP is part of the larger ClinGen Cardiovascular Domain Working Group.

### Specification of ACMG/AMP criteria

A core group of eleven FH VCEP members reviewed all criteria in the original ACMG/AMP guidelines and began to propose initial *LDLR*-specific modifications based on expert opinion and prior publications.^14^ Proposed modifications were discussed frequently through conference calls, emails, and several in-person meetings at international conferences, most notably the FH Foundation’s Global Summit (2016-2019) and European Atherosclerosis Society Congress (2016-2019), until consensus was reached. Proposed guidelines, in various iterations, were consistently evaluated in analyses using well-known variants as well as by reference to established literature. ClinGen’s Sequence Variant Interpretation (SVI) committee provided feedback and suggestions, which were incorporated in multiple rounds of revisions. Finalized criteria were ultimately voted upon and approved by all members of FH VCEP. Note that given differences in mechanisms of disease, prevalence and penetrance, it was decided that *APOB*- and *PCSK9*-specific guidelines will be completed separately.

### Validation and pilot testing

Following guideline approval from the SVI committee, a formalized pilot study of 54 *LDLR* variants was performed in the ClinGen Variant Curation Interface (VCI; https://curation.clinicalgenome.org/). The VCI is a publicly available, comprehensive resource that systematically facilitates individual and group-level curation activities in accordance with the ACMG/AMP guidelines.

Pilot study curations in the VCI were performed independently by two biocurators, followed by a review from two VCEP leadership members. Publicly available data used for curation were supplemented with internal case-level data from VCEP member laboratories. When applicable, internal laboratory data used in the classifications were uploaded and saved into the VCI. Following independent curation of the 54 *LDLR* pilot-variants, biocurators extracted the data from the VCI and sent it to the reviewers. Reviewers compared the individual ACMG/AMP criteria codes used for each variant, as well as the final classifications. Any discordance in the application of criteria codes or in the final classification for each variant was recorded. Discordances were resolved in discussion among the biocurators and reviewers. Final classifications were approved by the reviewers and were submitted to ClinVar under the FH VCEP affiliation. The ontology used for FH due to *LDLR* variation was MONDO:0007750, with semi-dominant inheritance (HP:0032113) and the reference sequence used for *LDLR* was NM_000527.5.

Rules for combining pathogenic and benign criteria codes follow the original ACMG/AMP scoring algorithm (Richards et al., 2015)^12^ (**Supplementary Table 1**).

## RESULTS AND DISCUSSION

### Summary of specifications

FH VCEP specifications for *LDLR* variant classification in FH are summarized in **Table 1**. The type of *LDLR-*specific alterations we made to the original ACMG/AMP criteria codes can be categorized into the following: 14 disease-specific/strength-level changes, 13 disease-specific changes, one strength-level change, and four clarification changes (based on recent ClinGen recommendations). Additionally, we found six criteria codes not applicable to *LDLR*. Key *LDLR* modifications include alteration of population data frequency thresholds, delineation of loss-of-function (LoF) variant types, functional study criteria specifications, co-segregation criteria specifications, and specific use and thresholds for *in silico* prediction tools.

**Table 1.**
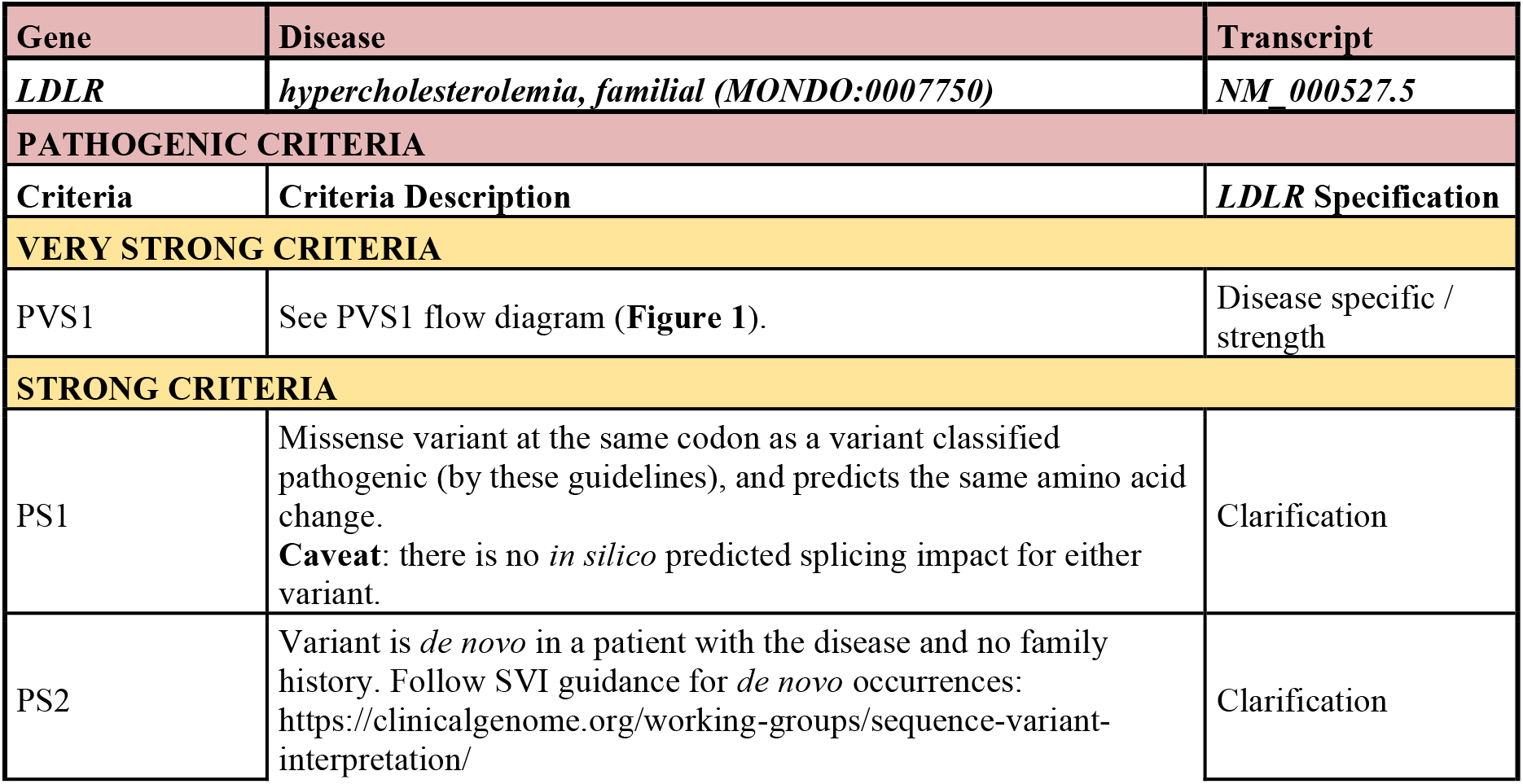
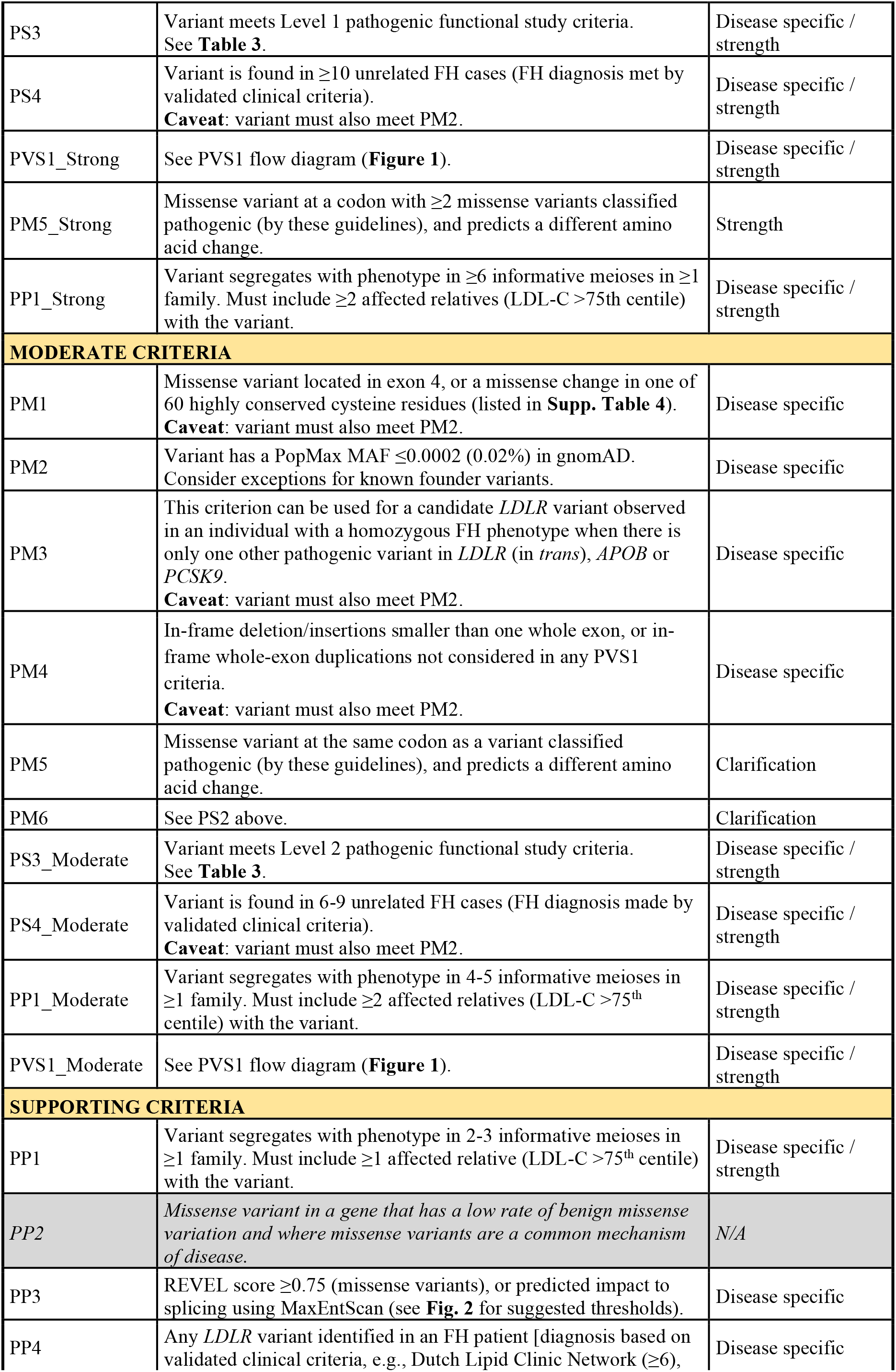
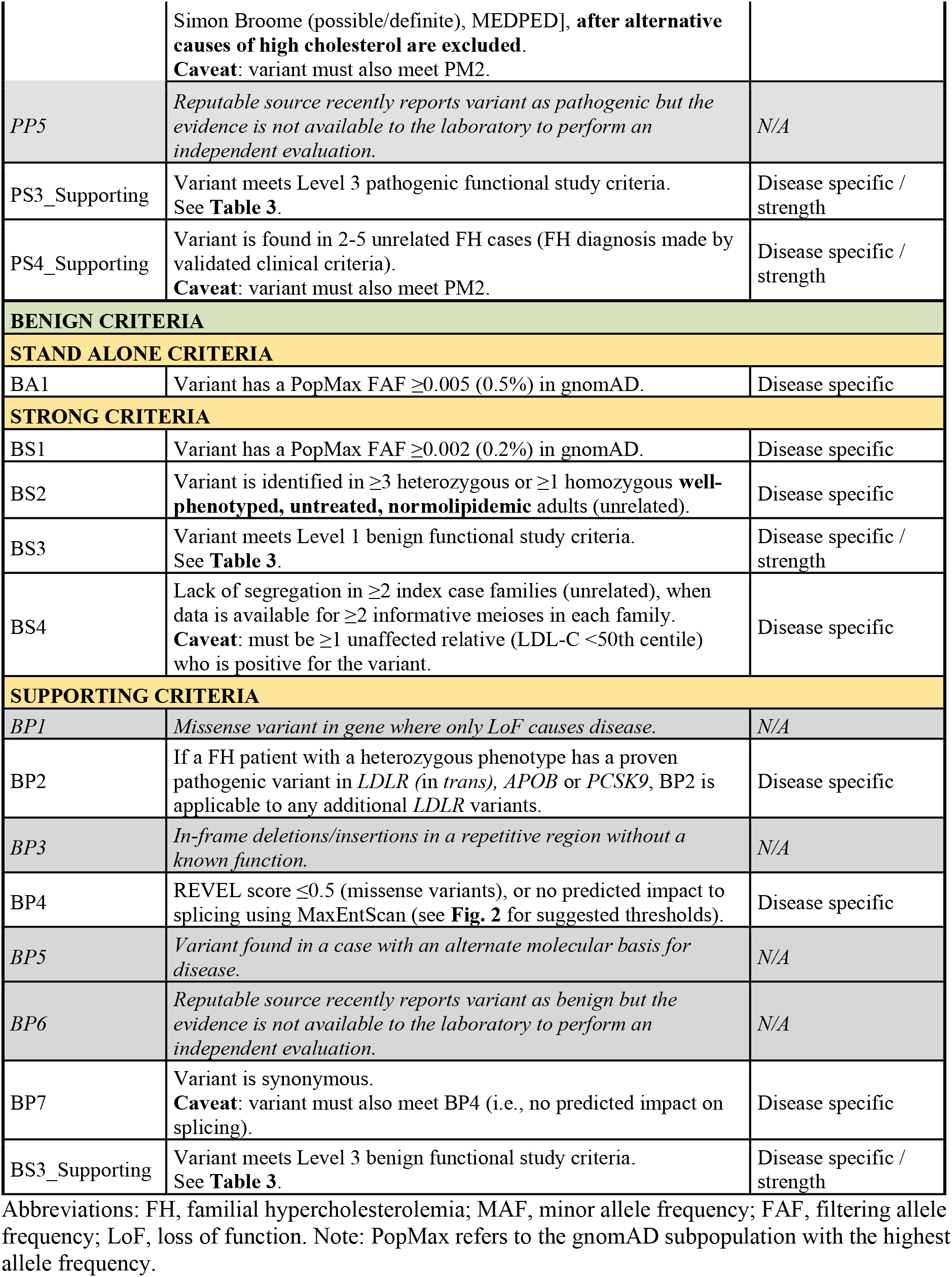
Summary of ACMG/AMP guideline specifications for *LDLR*.

### Population data (PM2, BA1, BS1)

The FH VCEP recommends using the gnomAD PopMax Filtering Allele Frequency (FAF) in evaluation of BA1 and BS1 codes^15^, while evaluation of PM2 should be performed using the PopMax Minor Allele Frequency (MAF). Frequency thresholds specific for *LDLR* variants in FH are displayed in **Table 2** and were calculated using the CardioDB metrics allele frequency web tool (https://www.cardiodb.org/allelefrequencyapp/) based on prevalence, penetrance, and allelic/genetic heterogeneity. Allele frequency thresholds were equal to FAF ≥0.005 (0.5%) for BA1, FAF ≥0.002 (0.2%) and <0.005 (0.5%) for BS1, and MAF ≤0.0002 (0.02%) for PM2. Note that if both exomes and genomes have a FAF/MAF value presented in gnomAD, consider the value corresponding to the higher number of alleles tested (i.e., higher total allele number). When evaluating whole-exon deletions and duplications, which are a relatively common pathogenic variant type in *LDLR* (∼10% of affected individuals),^16^ the gnomAD Structural Variant (SV) dataset should be queried.

**Table 2.** *LDLR*-specific population data frequency thresholds.

|  | **gnomAD**<br>**Frequency** | **Prevalence** | **Penetrance** | **Allelic Het.** | **Genetic Het.** |
| --- | --- | --- | --- | --- | --- |
| BA1 | PopMax FAF<br>$\geq 0.005$ (0.5%)<sup>a</sup> | 1/250 | 50% | 1.0 | 1.0 |
| BS1 | PopMax FAF<br>$\geq 0.002$ (0.2%) and<br>$< 0.005$ (0.5%) | 1/250 | 95% | 1.0 | 0.9 |
| PM2 | PopMax MAF<br>$\leq 0.0002$ (0.02%) | 1/250 | 95% | 0.1 | 0.9 |

Note: PopMax refers to the gnomAD subpopulation with the highest allele frequency. <sup>a</sup> BA1 metrics were equal to 0.4%; however, we conservatively increased the BA1 threshold to 0.5%. Abbreviations: FAF, filtering allele frequency; MAF, minor allele frequency; Het., heterogeneity.

It is important to keep in mind that both case and control gnomAD cohorts are expected to contain many individuals with FH, given FH is relatively common in the general population (1 in 250 individuals; or an estimated ∼34 million affected worldwide), and >90% of individuals are thought to be undiagnosed.^2,3^ Further, there are multiple cardiac case cohorts included in gnomAD, such as those from the Framingham Heart Study, Jackson Heart Study, Multi-Ethnic Study of Atherosclerosis, and Myocardial Infarction Genetics Consortium studies.

### Loss of function (PVS1) and in-frame indels (PM4)

*LDLR* satisfies ClinGen’s three requirements for applicability of PVS1.^17^ That is 1) it is a definitive gene for FH; 2) three or more LoF variants reach an ACMG/AMP classification of “Pathogenic” without PVS1 (see **Supplementary Table 2**); and 3) >10% of variants associated with the phenotype are LoF (across more than one exon). In fact, frameshift variants alone represent ∼20% of all unique FH-associated variants in ClinVar and are distributed throughout the gene.^11^

In accordance with the PVS1 flowchart outlined in Abou Tayoun et al., 2018,^17^ we have specified PVS1 (**Figure 1**) based on well-established evidence in *LDLR*. Notably, any stop codon amino-terminal of amino acid 830 (NM_000527.5; located in exon 17) has been shown to remove a region known to be critical to protein function (i.e., the NPXY sequence of the cytoplasmic tail, required for LDLR internalization).^18^ Note that alternative splicing of exons #1-18 from *LDLR*’s biologically relevant transcript is not known to occur. Therefore, PVS1 includes the following variants: 1) deletion of full gene; 2) deletion of single or multiple exons (exons 1-17) that lead to an out-of-frame consequence; 3) nonsense or frameshift variants causing a premature stop codon amino-terminal of amino acid 830 (NM_000527.5:p.Lys830); 4) variants in canonical +/− 1,2 GT/AG splice sites that predict a frameshift in exons 1-17; and 5) intragenic exon duplications proven to occur in tandem, that predict a frameshift in exons 1-17. PVS1_Strong includes: 1) deletion of single or multiple exons (exons 1-17) that do not predict a frameshift; 2) variants in canonical +/− 1,2 GT/AG splice sites that predict in-frame deletions in exons 1-17; and 3) intragenic exon duplications presumed to occur in tandem that predict frameshifts in exons 1-17. PVS1_Moderate includes: 1) variants in the initiation codon; 2) whole-exon deletion of exon 18; and 3) nonsense/frameshift variants carboxy-terminal of amino acid 830 (NM_000527.5:p.Lys830). Further, **Supplementary Table 3** provides information on the phase of *LDLR* exons for determining in-or out-of-frame consequences for applicable variants.

**Figure 1.**
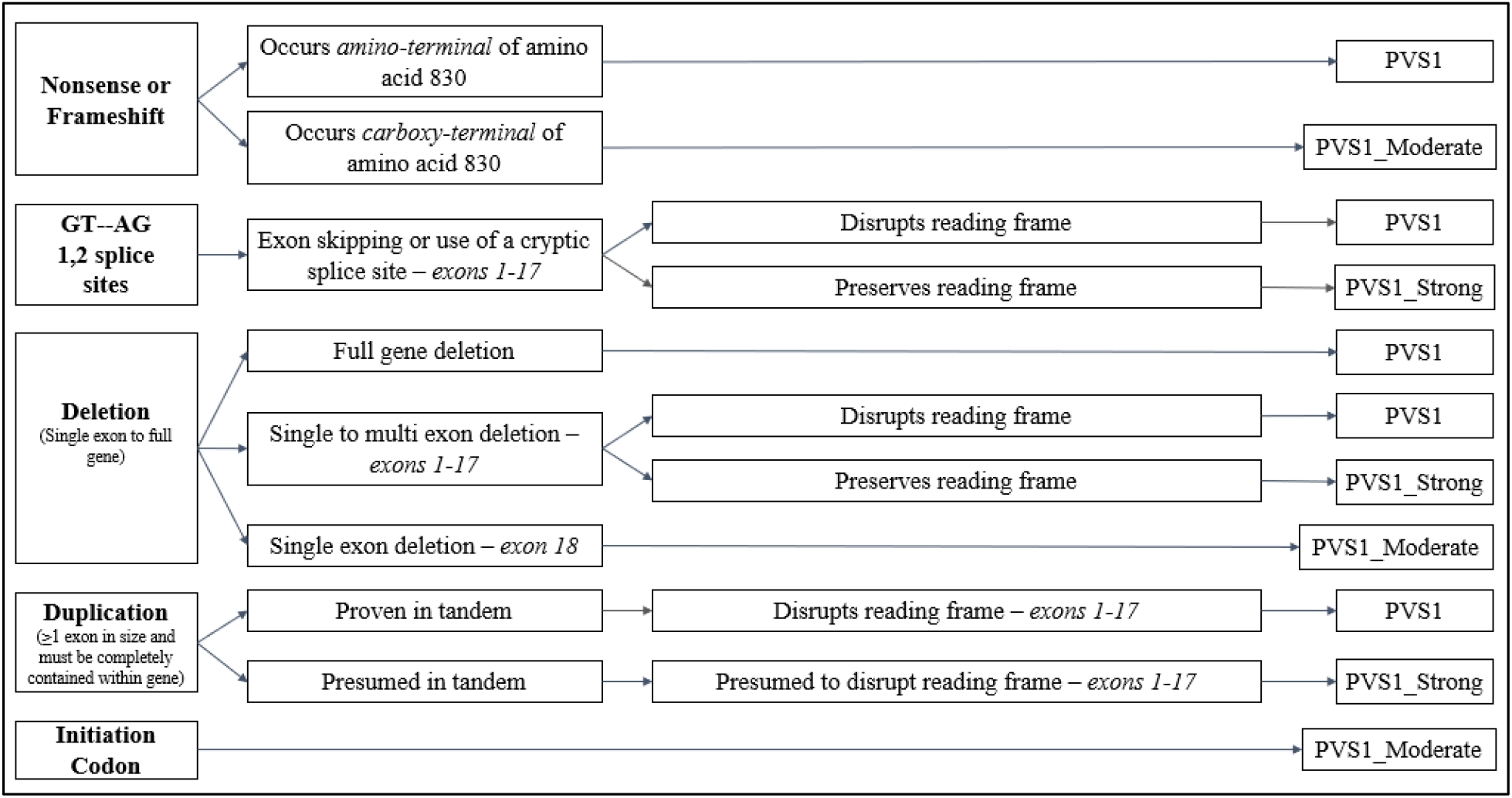
*LDLR*-specific recommendations for application of PVS1. Abbreviations: NMD, nonsense-mediated decay

In addition, in-frame deletions or insertions smaller than a whole exon, or in-frame whole-exon duplications not considered in the PVS1 criteria are applicable to PM4, if they also meet PM2 (i.e., MAF ≤0.0002 (0.02%)).

### Experimental studies (PS3, BS3)

Following the SVI recommendations for application of functional studies codes PS3/BS3,^19^ we have defined the mechanism of disease, evaluated the applicability of classes of assays used in the field, and evaluated individual instances of assays, in determining appropriate strength levels.

In summary, LDLR is expressed at the cell surface, where it binds circulating plasma LDL particles. The LDLR-LDL (receptor-ligand) complex is internalized at clathrin-coated pits via receptor-mediated endocytosis. Once internalized (as part of the endosome), acidic conditions mediate release of the LDL ligand from its receptor, and the receptor is recycled back to the cell surface where it can repeat this process; a single LDLR protein can be recycled ≥100 times.^20^ Pathogenic variants may induce a loss of function at any part of the LDLR cycle,^21^ disrupting LDLR activity, and leading to FH due to an inability to effectively clear LDL-C from the bloodstream. The most reliable functional assays are adapted from the Nobel Prize winning work of Drs. Michael Brown and Joseph Goldstein^22^ and allow the characterization of the whole LDLR cycle, which can be evaluated sufficiently at three key steps: 1) LDLR expression/biosynthesis, 2) LDL particle binding, and 3) LDL internalization. Such assays compare LDLR activity in wild-type cells against cells harboring a specific variant and are currently performed by flow cytometry with fluorescently-labeled LDL (commercially available or isolated from a wild-type individual) in 1) heterologous cells (with no endogenous LDLR) transfected with a mutant plasmid or 2) patient cells (fibroblasts, lymphocytes or lymphoblasts). Older studies using radioactively-labelled LDL (^125^I-LDL) (e.g., Hobbs et al., 1992)^23^ are also valid if they use these same cell types. A more in-depth analysis of the rationale and methodologies of LDLR functional assays are presented in Bourbon, Alves, & Sijbrands, 2017.^24^

We have determined three different strength “levels” of LDLR functional assays (see **Table 3**), based on the appropriateness of the methodology. Level 1 studies (set at PS3/BS3) are the most reliable; these include flow cytometry assays which evaluate the whole LDLR cycle (i.e., LDLR expression/biosynthesis, LDL binding and LDLR-LDL internalization) performed in heterologous cells (with no endogenous LDLR) transfected with a mutant plasmid. Using heterologous cells with site-directed mutagenesis ensures that the assay is variant-specific. **Supplementary Figure 1** demonstrates thresholds and controls used to validate flow cytometry assays in heterologous cells. Level 2 and Level 3 studies (set at PS3_Moderate, PS3_Supporting/BS3_Supporting) represent additional techniques that allow for evaluation of only part of the LDLR cycle, or which use less robust cells/materials. It is important to note that when using patient cells, DNA sequence analysis of *LDLR* should indicate the assay is variant-specific (i.e., no other candidate variants identified in *LDLR*, including whole-exon deletions/duplications). Although the historical Brown and Goldstein LDLR activity assays using patient cells were thoughtfully designed to be gene-specific (for e.g., APOB-containing LDL particles used are always wild-type and LDLR is overexpressed in the cultured cells), patient-specific genetic factors may still modify outcomes. Lastly, studies in compound heterozygous patient cells are not considered as valid functional assays since it is difficult to delineate the individual effect of each variant.

**Table 3.** PS3/BS3 functional study criteria specifications for *LDLR*.

| **Pathogenic** |  |
| --- | --- |
| PS3<br>(Level 1) | (1) Study of the *whole* LDLR cycle (LDLR expression/biosynthesis, LDL binding, and LDL internalization) performed in heterologous cells (with no endogenous LDLR) transfected with a mutant plasmid. Assay result of <70% of wild-type activity in either expression/biosynthesis, binding OR internalization. |
| PS3_Moderate<br>(Level 2) | (1) Study of a) only *part* of the LDLR cycle following Level 1 methodology, or b) *whole or part* of the LDLR cycle in true homozygous patient cells. A variant with assay results of <70% of wild type activity in either LDLR expression/biosynthesis, LDL binding OR internalization.<br>(2) RNA studies, using RNA extracted from heterozygous or true homozygous patient cells, where aberrant transcript is confirmed by sequencing and is quantified as >25% of total transcript from heterozygous cells or 50% of total transcript from homozygous cells.<br>(3) Variants with two or more Level 3 functional studies (must be different assays); or any Level 3 functional study #1-4 performed by two or more independent labs with concordant results. |
| PS3_Supporting<br>(Level 3) | (1) Study of LDLR cycle (*whole or part*) in heterozygous patient cells, with assay results of <85% of wild-type activity in either LDLR expression/biosynthesis, LDL binding OR internalization.<br>(2) Luciferase studies with transcription levels of <50% compared to wild-type (applicable to 5'UTR/promoter variants).<br>(3) Minigene splicing assays with <10% wild-type transcript present where an aberrant transcript from the candidate variant is confirmed by sequencing.<br>(4) High-throughput assays, which include alternative microscopy assays (e.g., Thormaehlen et al., 2015), Multiplex Assays of Variant Effect (MAVE) (e.g., Weile & Roth, 2018) and deep mutational scanning assays, can be considered here, only if assay has been validated with a minimum of four pathogenic and four benign variant controls in LDLR. *Note: % activity thresholds will be defined by the FH VCEP as more data becomes available.<br>(5) RNA studies, using RNA extracted from heterozygous or homozygous patient cells, with aberrant transcript confirmed by sequencing (but without transcript quantification). |
| **Benign** |  |
| BS3<br>(Level 1) | (1) Study of the *whole* LDLR cycle (LDLR expression/biosynthesis, LDL binding, and LDL internalization) performed in heterologous cells (with no endogenous LDLR) transfected with a mutant plasmid. Assay result of >90% of wild-type activity in expression/biosynthesis, binding AND internalization.<br>Note: studies of only *part* of the LDLR cycle are not eligible for BS3 or BS3_Supporting. |
| BS3_Supporting<br>(Level 3) | (1) Study of the *whole* LDLR cycle in a) true homozygous patient cells, with assay result of >90% of wild-type activity in biosynthesis, binding AND internalization; or in b) heterozygous patient cells with assay result of >95% of wild-type activity in biosynthesis, binding AND internalization.<br>(2) Luciferase studies with transcription levels of >90% when compared to wild-type (applicable to 5'UTR/promoter variants).<br>(3) RNA studies, using RNA extracted from heterozygous or true homozygous patient cells, with a) aberrant transcripts quantification, where aberrant transcript is <10% of total transcript OR b) without transcript quantification where no aberrant transcript is confirmed by sequencing.<br>(4) Minigene splicing assay where only wild-type transcript is present and confirmed by sequencing.<br>(5) High-throughput assays as defined above; only applicable when assay can indicate the *whole* LDLR cycle (LDLR expression/biosynthesis, LDL binding AND internalization) is unaffected. |

Note: functional assays performed in compound heterozygous patient cells are not considered applicable in PS3/BS3 criteria since it is difficult to delineate the individual effect of each variant.

### Hotspot/well-established functional domains (PM1)

*LDLR* exon 4 is considered a mutational hot spot for missense variants in a well-established functional domain critical to protein function, since it encodes LDLR type A repeats 3, 4 and 5, which compose the well-established ligand (LDL) binding domain;^25^ exon 4 also has the highest number of FH-associated variants per nucleotide with no variants proven benign by functional studies.^14^ In addition, *LDLR* contains 60 highly conserved cysteine residues (located throughout exons 2-8 and 14) critical to protein function; these 60 cysteine residues are involved in disulfide bond formation, essential for proper protein folding.^26,27^ Thus, PM1 is applicable to any missense change in the amino acids of exon 4 (NM_000527.5:c.314-694 or p.105-232) that are also rare (i.e., PM2 criterion is met), or to any missense change in the 60 highly conserved cysteines, which we have listed in **Supplementary Table 4**.

### Observed in healthy adults (BS2)

Pathogenic variants in *LDLR* are known to be highly penetrant, where “affected” status is typically identifiable as early as childhood^28^ through a simple and routine laboratory measure of plasma LDL-C level. Therefore, we have determined BS2 is applicable for *LDLR* variants identified in ≥3 heterozygous or ≥1 true homozygous well-phenotyped, normolipidemic, untreated, unrelated adults. At a minimum, “well-phenotyped” refers to LDL-C measurements taken over multiple time points (≥2), with consistent results. Individuals considered in BS2 should not be taking any lipid-lowering therapy near the time of measurement and should have an LDL-C level below the ethnic and country-specific 50th centile (adjusted for age and sex) (e.g., Starr et al. 2008).^29^ Because lipid-lowering therapies (e.g., statins) are among the most widely prescribed medications in the general population, and neither medication status nor LDL-C level are typically available in commonly used, publicly available resources such as gnomAD or ExAC, such resources must not be used for evaluation of BS2 in FH. Rather, we recommend evaluation of BS2 in well-phenotyped normolipidemic cohorts only, which are likely to be more available in internal laboratory settings. It is important to follow these caveats closely, given BS2 is a strong-level criterion.

### Specificity of phenotype (PP4) and case-control data (PS4)

There are a variety of validated clinical diagnostic criteria used for FH, which include the Dutch Lipid Clinic Network (DLCN) criteria,^30^ Simon Broome criteria,^31^ the United States MEDPED criteria,^32^ and other country-specific criteria. We have determined that PP4 is applicable to any rare (i.e., PM2 criterion is met) *LDLR* variant identified in a patient with a diagnosis of FH based on any validated clinical criteria; examples include a DLCN score ≥6, Simon Broome score of “possible” or “definite” FH, or a MEDPED diagnostic score of “FH”. Because 1) all validated clinical criteria require extreme LDL-C levels to be present in the patient together with a family history positive for high LDL-C and/or premature coronary heart disease, and 2) the *LDLR* gene is specific for FH (>90% of cases), we believe strongly in the appropriateness of PP4 for *LDLR*. However, in any case, PP4 is applicable only after alternative causes of high LDL-C are excluded. Alternative causes for high LDL-C are reviewed in Sturm et al., 2018^7^ and include polygenic dyslipidemia, elevated lipoprotein(a) [Lp(a)], nephrotic syndrome, obstructive liver disease, hypothyroidism, FH due to *PCSK9, APOB*, or *APOE* variants, or FH phenocopies due to bi-allelic variants in *LDLRAP1, LIPA*, or *ABCG5/8*.

For the case-control criterion PS4, different strength levels may be applied depending on the number of unrelated FH cases with the rare variant. PS4 is applicable if the variant is found in ≥10 unrelated FH cases (FH diagnosis met using validated clinical criteria); PS4_Moderate is applicable if found in 6-9 unrelated FH cases; and PS4_Supporting is applicable if found in 2-5 unrelated FH cases. Note that in applying PS4-level criteria, the variant must also meet PM2 (i.e., MAF ≤0.0002 (0.02%)).

### Segregation data (PP1, BS4)

We have determined three strength levels for application of PP1 depending on the number of families/individuals studied. PP1_Strong is applicable when there is co-segregation of the variant with affected status in ≥6 informative meioses; PP1_Moderate when in 4-5 informative meioses; and PP1_Supporting when in 2-3 informative meioses. Index cases should not be counted as positive cases for co-segregation results. When the same variant is identified in more than one family, data can be added to reach stronger evidence levels. **Supplementary Figure 2** shows a typical example of co-segregation in a pedigree, with an explanation on informative meiosis for a FH-associated variant. Note that when an index case presents with a heterozygous FH phenotype and the hypercholesterolemia is associated with one branch of the family, individuals from the other branch should not be considered for co-segregation analysis. BS4 is applicable when there is lack of co-segregation in ≥2 index case families (unrelated) and there is data on ≥2 informative meioses in each family. When applying BS4 there should be at least one instance where an unaffected family member carries the variant (i.e., genotype-positive, phenotype-negative).

For co-segregation analysis we consider an affected individual as one with an untreated total cholesterol (TC) or LDL-C level above the 75th centile adjusted for age and sex. Each country/region should preferably use their TC and LDL-C centile charts. Given the widespread use of lipid-lowering therapies in the general population, untreated TC or LDL-C measurements may not be obtainable for some individuals under consideration for PP1/BS4. For those with only known *treated* TC or LDL-C levels, several imputation factors may be applied for an estimation of untreated measurements; namely by specific medication and dose (preferred)^33^, or by the more general 0.8 and 0.7 correction factors corresponding to an estimated 20% TC and 30% LDL-C reduction on treatment, respectively.^34^ Unaffected family members should have ‘untreated’ TC and LDL-C below the 50^th^ centile adjusted for age and sex.

It is important to consider both affected and unaffected individuals when evaluating co-segregation. Alternative causes of high TC or LDL-C values, such as those described above, should be considered carefully given their ability to explain instances of hypercholesterolemia in genotype-negative family members. It is important to note that cholesterol concentrations are influenced by the co-inheritance of common variants of small effect; Trinder et al. have recently demonstrated that individuals who have a LDL-C polygenic risk score in the lowest decile have LDL-C concentrations considerably lower than those in the highest decile (3.61 mmol/L versus 4.37 mmol/L, respectively)^35^. Lastly, be aware that although rare, FH patients with a pathogenic *LDLR* variant could also be positive for a rare monogenic cholesterol-lowering variant (possible in *APOB* or *PCSK9* genes for example), as has been described in Emi et al., 1991^36^ and Motazacker et al., 2012.^37^ If identifiable, these individuals should not be considered for co-segregation analysis.

### *In silico* prediction (PP3, BP4)

For *in silico* classification of missense variants in *LDLR* we suggest the use of REVEL, an ensemble method for pathogenicity prediction that combines predictions from 13 individual commonly used computational tools: MutPred, FATHMM, VEST, Poly-Phen, SIFT, PROVEAN, MutationAssessor, MutationTaster, LRT, GERP, SiPhy, phyloP, and phastCons.^38^ REVEL has proved to be consistent and to outperform existing tools in distinguishing pathogenic from rare neutral variants. REVEL was also selected because of its accessibility; REVEL scores are pre-computed and automatically displayed in the ClinGen VCI (under the ‘Variant Type’ tab), or are available for download (LDLR: https://rothsj06.u.hpc.mssm.edu/revel/revel_segments/revel_chrom_19_009082971-013246689.csv.zip). Use of a single meta-predictor such as REVEL will eliminate discrepancies in which programs are used by curators, which transcripts are selected, how many tools should be concordant and what specificities to account for when manually performing *in silico* analysis. To determine *LDLR*-specific score thresholds for PP3/BP4 we evaluated REVEL scores for *LDLR* missense variants with 1) Pathogenic/Likely pathogenic or Benign/Likely Benign classifications in ClinVar, 2) damaging or neutral results according to *LDLR*-specific PS3/BS3 functional study evidence, and 3) concordant *in silico* results for Poly-Phen, SIFT, PROVEAN and MutationTaster (**Supplementary Figure 3**). Using the PP3 threshold of ≥0.75 (as defined in Ioannidis et al., 2016)^38^, the vast majority of variants with an association of ‘pathogenic’ using any comparison (as above) have REVEL scores above this threshold. Using the BP4 threshold of ≤0.5, ∼half of variants with an association of ‘benign’ from ClinVar classifications or other *in silico* predictors have REVEL scores below this threshold. Therefore, we recommend a REVEL score ≥0.75 as supportive evidence of pathogenicity (PP3), and a REVEL score ≤0.5 as supportive evidence of benign (BP4).

For *in silico* prediction of splicing effects, we recommend evaluation only if no functional data is available; furthermore, variants already considered in PVS1 (or modified strength) (i.e., those which are in the canonical +/− 1,2 GT/AG splice sites) should not be further considered in PP3/BP4. We suggest the use of MaxEntScan (MES)^39^ which is highly reputable and publicly available. We have defined distinct thresholds for MES depending on the variant location, as described in **Figure 2**.

**Figure 2.**
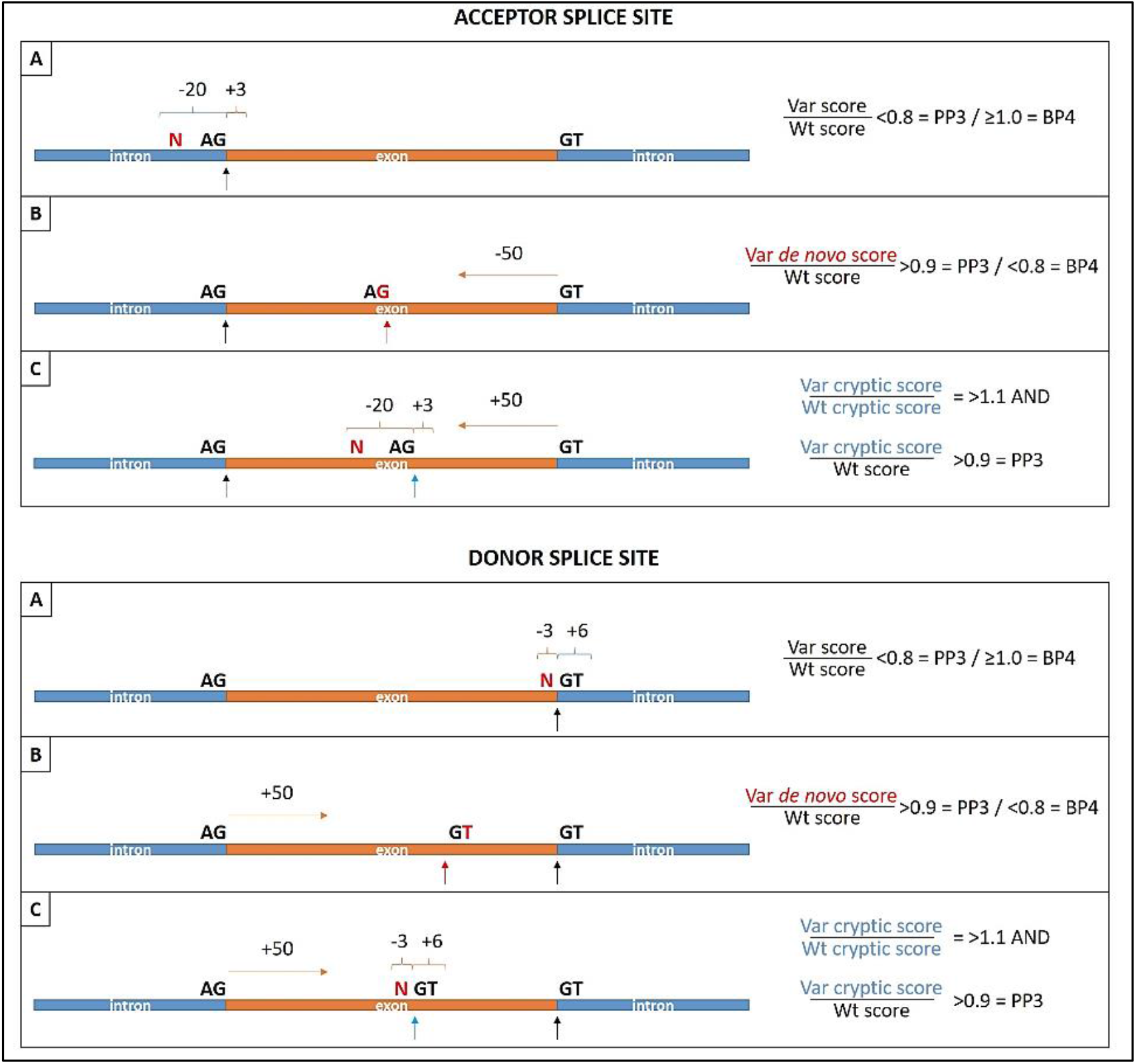
FH VCEP suggestions for evaluating splicing effects using MaxEntScan (MES) dependent on variant location A, B, or C. (**A**) Variant is located at −20 to +3 bases related to the authentic acceptor splice site or at −3 to +6 related to the authentic donor splice site: A result of authentic splice site strength variant/wild-type score <0.8 is supportive evidence of pathogenicity (PP3), while a score ≥1.0 is supportive evidence of benign (BP4). (**B**) Variant creates *de novo* acceptor splice site, which is at least 50 bases upstream of the authentic donor splice site, or *de novo* donor splice site, which is at least 50 bases downstream of the authentic acceptor splice site: A result of *de novo* splice site strength variant/authentic wild-type score in >0.9 is applicable to PP3, while a score <0.8 is applicable to BP4. (**C**) Variant is located at −20 to +3 bases relative to an intra-exonic AG dinucleotide, which is at least 50 bases upstream of the authentic donor splice site, or at −3 to +6 bases relative to an intra-exonic GT dinucleotide, which is at least 50 bases downstream of the authentic acceptor splice site: Results of both variant cryptic/wild-type cryptic score in >1.1 and cryptic acceptor/authentic acceptor score or cryptic donor/authentic donor score in >0.9 is applicable to PP3. Note: BP4 is applicable to exonic variants outside of the 50 base limits detailed above, given the unlikelihood of such variants to impact splicing in *LDLR*. Abbreviations: Var, variant; Wt, wild-type.

Lastly, if both missense and splicing prediction are applicable, only one prediction of a damaging effect is sufficient in applying PP3; however, both need to predict a neutral effect in applying BP4.

### Other variants in the same codon (PS1, PM5)

When there are other described variants in the same codon as a missense variant being classified, PS1 is applicable if at least one missense variant has a classification of pathogenic (classified using these *LDLR*-specific guidelines), and the variant predicts the same amino acid change. PM5 is applicable if there is one pathogenic missense variant that predicts a different amino acid in the same codon. Lastly, PM5_Strong is applicable in the same context if there are ≥2 pathogenic missense variants which predict different amino acids in the same codon. Note that for these codes to be applied, the curated variant(s) should 1) not already be considered in PM1 (hotspot/well-established functional domain), and 2) have an *in silico* predicted splicing impact of benign. Combining PS1/PM5 with PM1 can be considered “double-counting”, i.e., evaluating a variant under a similar premise twice, while investigating potential splicing impact provides greater confidence that pathogenicity is related to a predicted altered amino acid rather than creation of a *de novo* splice site or activation of cryptic splice site.

### Allele data (*cis/trans*) (PM3, BP2)

*LDLR* variants show a semi-dominant pattern of inheritance on plasma cholesterol concentration, such that the phenotypes in homozygous or compound heterozygous patients (i.e., those with bi-allelic variants) are significantly more severe than in heterozygotes (i.e., those with mono-allelic variants). Because of this, both PM3 and BP2 criteria (observed in *trans* with a pathogenic variant) can be used when case-level data are available for individuals with more than one FH-associated variant. PM3 is applicable when a candidate *LDLR* variant is identified in a patient with a clear homozygous or compound heterozygous FH phenotype (defined here as untreated LDL-C ≥13 mmol/L or ≥500 mg/dl), who has an additional known pathogenic variant in *LDLR* (in *trans*), *APOB*, or *PCSK9*. The candidate variant must also meet PM2 (i.e., MAF ≤0.0002 (0.02%)). PM3 must not be used if *cis/trans* status in *LDLR* has not been established. BP2 is applicable to any additional *LDLR* variants identified in a patient with a clear heterozygous FH phenotype (defined here as untreated, elevated LDL-C that is <8 mmol/L or <310 mg/dl, in adults) who already has a known pathogenic variant in *LDLR (*in *trans), APOB*, or *PCSK9*.

For both PM3 and BP2, known pathogenic variants in *LDLR* must be classified as pathogenic according to these guidelines, while known pathogenic variants in *APOB* or *PCSK9* should be formally assessed by general ACMG/AMP guidelines until these gene-specific guidelines have been established.

### *De novo* occurrence (PS2, PM6)

The FH VCEP recommends following the SVI recommendations for PM6 and PS2, which can be found at https://clinicalgenome.org/working-groups/sequence-variant-interpretation. These recommendations evaluate PM6/PS2 based on a points system centered around three parameters: confirmed versus assumed status, phenotypic consistency, and number of *de novo* observations. Although data to address *de novo* occurrence in *LDLR* directly are lacking, we have no evidence to suggest that this a common feature in FH, given that to date, only one member of FH VCEP has observed a *de novo* occurrence in their clinical practice, and to the best of our knowledge, such cases have only been reported once in the literature.^40^ However, they are of course possible, and should be considered.

### Criteria not applicable (BP1, PP2, BP3, BP6, PP5, BP5)

BP1 (missense variant in a gene for which primarily truncating variants are known to cause disease) is not applicable, since the majority of FH-associated *LDLR* variants are missense variants. Following SVI counsel regarding PP2 (missense variant in a gene that has a low rate of benign missense variation and where missense variants are a common mechanism of disease), PP2 is not applicable, on the basis of a low z-score = 0.12 for *LDLR* in the gnomAD missense constraint table. BP3 (in-frame deletions/insertions in a repetitive region without a known function) is not applicable, given that there are no regions in *LDLR* without a known function. BP6 and PP5 (variant previously classified by a reputable source) have been advised by ClinGen not to be used, so are not applicable. Lastly, the FH VCEP has decided to remove BP5 (alternative mechanism for disease), given that this premise is already evaluated in our specifications for BP2.

### Pilot study

The pilot study of the final specifications was done on 54 *LDLR* variants in the ClinGen VCI. Variants, listed in **Supplementary Table 5**, were chosen to reflect *LDLR* variant variability and included one multi-exon deletion, 40 missense, seven intronic/splicing, four nonsense and two synonymous variants. A total of nine institutions, which included six clinical and three research laboratories, provided internal case-level data (via standardized template), supplementing classifications for 42 of 54 variants. For the remaining variants, and whenever necessary, data from published literature was used to count number of cases, and to evaluate co-segregation and functional evidence.

Preliminary results had complete agreement (in both classification and individual criteria used) for 16 variants, agreement in classifications but not in each criterion used for 27 variants, and discrepancies in both classification and criteria used for the remaining 11 variants. Differences in classification were eight counts of Likely pathogenic versus VUS, two of Likely pathogenic versus Pathogenic, and one of Benign versus VUS. A careful review of the discrepancies determined that most resulted from extracting different gnomAD MAF/FAF data from the VCI, or from differences in applying PS4_moderate versus PS4_supporting due to slight differences in case counts. Minor refinements were added to the guidelines to address these discrepancies: we clarified the use of MAF/FAF (exomes versus genomes feature) in gnomAD and designed a more efficient template for tracking case-level data. Consequently, finalized pilot results had complete agreement in both classification and criteria used for all 54 variants and represented six Benign, two Likely benign, 18 VUS, 15 Likely pathogenic and 13 Pathogenic variants. The number of times each criterion was used is represented in **Supplementary Figure 4**. Reviewers approved the final classifications which are now live in ClinVar under the FH VCEP affiliation.

## LIMITATIONS

There are multiple criteria specifications that require diagnostic information and case-level data, which if not readily available, may limit the classification of variants. For instance, PS4 and PP4 criteria require that cases considered are clinically diagnosed with FH based on validated clinical criteria. PP1-level, PS2/PM6, PM3, BS2, BS4, and BP2 criteria require that further case-level data are available, including LDL-C measurements, genetic results, family history, and medication status. However, although this information may be difficult to ascertain in some settings, it is necessary to apply these criteria correctly. Whenever possible, we encourage curators to actively seek this information if it is not initially available. As the FH VCEP works toward classifying all ∼2,300 *LDLR* variants currently in the ClinVar database using the guidelines presented here, we are hopeful we can overcome some of these limitations through internal data sharing efforts. We encourage any laboratory with internal data on *LDLR* variants in FH patients to upload these data in ClinVar, as these data can have a major impact on the proper classification of variants.

## CONCLUSIONS AND FUTURE DIRECTIONS

Here, the FH VCEP presents consensus recommendations for *LDLR* variant classification. Application of these guidelines will provide evidence-based, standardized classification of *LDLR* variants for use in clinical diagnostics and research. Future directions include sustained variant curation, with the aim of classifying all ∼2,300 unique *LDLR* variants in ClinVar at the ‘3-star’ status. It is noteworthy that in 2018 the ClinGen Variant Curation Expert Panel protocol was recognized by the U.S. Food and Drug Administration (FDA), whereby variant classifications with 3-star status in ClinVar are now associated with an FDA-recognized tag. In the future, we expect this may have implications for obtaining insurance coverage for certain medications, for enrolment in certain clinical trials or research studies, or in the feedback of incidental findings from whole-exome or whole-genome sequencing, such as in the UK 100,000 genomes project. Given these possible implications, we will prioritize the classification of *LDLR* variants with the greatest potential impact, such as those that are LoF variant types, those with many and/or conflicting submissions currently in ClinVar, or those known to be on clinically available arrays/panels. We are hopeful that the FH VCEP classification of all ∼2,300 *LDLR* variants is completed within the next four years; we also plan to review all classifications on a two-year basis to ensure that recently emerging data are considered.

Finally, please be aware that the *LDLR*-specific guidelines presented here are subject to change in response to emerging data and newly available resources, which will continually influence the evolving nature of variant classification methodology, both specific to *LDLR* and also more broadly throughout the clinical genetics community. For this reason, please refer to the FH VCEP page in the ClinGen website (https://clinicalgenome.org/affiliation/50004) for the most currently accepted version.

## Data Availability

All data generated or analyzed during this study are included in this article (and its supplementary information files).

## ACKNOWLEDGEMENTS

ClinGen is primarily funded by the National Human Genome Research Institute (NHGRI), through the following three grants: U41HG006834, U41HG009649, U41HG009650. ClinGen also receives support for content curation from the Eunice Kennedy Shriver National Institute of Child Health and Human Development (NICHD), through the following three grants: U24HD093483, U24HD093486, U24HD093487. The content is solely the responsibility of the authors and does not necessarily represent the official views of the National Institutes of Health. JRC acknowledges her PhD fellowship funded by the Science and Technology Foundation (SFRH/BD/108503/2015). LT and TF are supported by the Ministry of Health of the Czech Republic, grant NU20-02-00261. SEH is an Emeritus British Heart Foundation Professor and is funded by PG08/008, and by the National Institute for Health Research University College London Hospitals Biomedical Research Centre. MT is supported by a Vanier Canada Graduate Scholarship. LRB is a Michael Smith Foundation for Health Research Scholar and a Canada Research Chair in Precision Cardiovascular Disease Prevention. RAH is supported by the Jacob J. Wolfe Distinguished Medical Research Chair, the Edith Schulich Vinet Canada Research Chair in Human Genetics, the Martha G. Blackburn Chair in Cardiovascular Research, and operating grants from the Canadian Institutes of Health Research (Foundation Grant) and the Heart and Stroke Foundation of Ontario (G-18-0022147). JWK is supported by the NIH through grants P30DK116074 (to the Stanford Diabetes Research Center), R01 DK116750, R01 DK120565, R01 DK106236; and by the American Diabetes Association (grant #1-19-JDF-108). Lastly, the FH VCEP would like to thank Drs. Steven Harrison, Leslie Biesecker, Heidi Rehm and the additional members from the ClinGen SVI for their feedback and guidance in establishing these guidelines.

