## Supplemental Tables and Figures for "The Clinical Genome Resource (ClinGen) Familial Hypercholesterolemia Variant Curation Expert Panel consensus guidelines for *LDLR* variant classification"

### SUPPLEMENTARY TABLES AND FIGURES

**Supplementary Table 1**. Rules for combining pathogenic and benign criteria in ACMG/AMP guideline specifications for *LDLR*.

| | | |
|---|---|---|
| **PATHOGENIC** | | |
| 1 Very Strong AND | 1 or more Strong | |
| | 2 or more Moderate | |
| | 1 Moderate AND | 1 Supporting |
| | 2 or more Supporting | |
| ≥2 Strong | | |
| 1 Strong AND | 3 or more Moderate | |
| | 2 Moderate AND | 2 or more Supporting |
| | 1 Moderate AND | 4 or more Supporting |
| **LIKELY PATHOGENIC** | | |
| 1 Very Strong AND | 1 Moderate | |
| 1 Strong AND | 1-2 Moderate | |
| | 2 or more Supporting | |
| 3 or more Moderate | | |
| 2 Moderate AND | 2 or more Supporting | |
| 1 Moderate AND | 4 or more Supporting | |
| **BENIGN** | | |
| 1 Stand Alone | | |
| 2 or more Strong | | |
| **LIKELY BENIGN** | | |
| 1 Strong AND | 1 Supporting | |
| 2 or more Supporting | | |
| **Variant of Uncertain Significance (VUS)** | | |
| Criteria shown above are not met OR the criteria for pathogenic and benign are contradictory | | |

Adapted from Richards et al., 2015[1]; no changes to original scoring algorithm.

**Supplementary Table 2**. *LDLR* loss-of-function variants that reach an ACMG/AMP classification of “Pathogenic” without the application of PVS1.

| ***LDLR* variant** | **Applicable criteria*** | **Sum of criteria** |
|---|---|---|
| c.313+1G>A | (PVS1_Strong); PS4; PP1_Strong; PM2; PS3_Moderate; PP4 | 2 Strong, 2 Moderate and 1 Supporting |
| c.564C>G (p.Tyr188Ter) | (PVS1); PP1_Strong; PS3_Moderate; PM2; PS4_Supporting; PP4 | 1 Strong, 2 Moderate and 2 Supporting |
| c.2140+1G>A | (PVS1); PP1_Strong; PM2; PS4_Moderate; PS3_Supporting; PP4 | 1 Strong, 2 Moderate and 2 Supporting |

*The criteria in parentheses in this column were not applied.

**Supplementary Table 3**. *LDLR* exon information.

| Exon No. | Start (g.) | Stop (g.) | Start (c.) | Stop (c.) | Length | Start Phase | End Phase |
|---|---|---|---|---|---|---|---|
| 1 | 11089463 | 11089615 | -86 | 67 | 153 | - | 1 |
| 2 | 11100223 | 11100345 | 68 | 190 | 123 | 1 | 1 |
| 3 | 11102664 | 11102786 | 191 | 313 | 123 | 1 | 1 |
| 4 | 11105220 | 11105600 | 314 | 694 | 381 | 1 | 1 |
| 5 | 11106565 | 11106687 | 695 | 817 | 123 | 1 | 1 |
| 6 | 11107392 | 11107514 | 818 | 940 | 123 | 1 | 1 |
| 7 | 11110652 | 11110771 | 941 | 1060 | 120 | 1 | 1 |
| 8 | 11111514 | 11111639 | 1061 | 1186 | 126 | 1 | 1 |
| 9 | 11113278 | 11113449 | 1187 | 1358 | 172 | 1 | 2 |
| 10 | 11113535 | 11113762 | 1359 | 1586 | 228 | 2 | 2 |
| 11 | 11116094 | 11116212 | 1587 | 1705 | 119 | 2 | 1 |
| 12 | 11116859 | 11116998 | 1706 | 1845 | 140 | 1 | 0 |
| 13 | 11120092 | 11120233 | 1846 | 1987 | 142 | 0 | 1 |
| 14 | 11120370 | 11120522 | 1988 | 2140 | 153 | 1 | 1 |
| 15 | 11123174 | 11123344 | 2141 | 2311 | 171 | 1 | 1 |
| 16 | 11128008 | 11128085 | 2312 | 2389 | 78 | 1 | 1 |
| 17 | 11129513 | 11129670 | 2390 | 2547 | 158 | 1 | 0 |
| 18 | 11131281 | 11133820 | 2548 | 2583 | 35 | 0 | - |

Phase: the position of an exon/intron boundary within a codon. A phase of zero means the boundary falls between codons, one means between the first and second base and two means between the second and third base. Genomic (g.) cooridinates correspond to reference sequence NC_000019.9, and coding (c.) cooridinates correspond to *LDLR* transcript NM_000527.5.

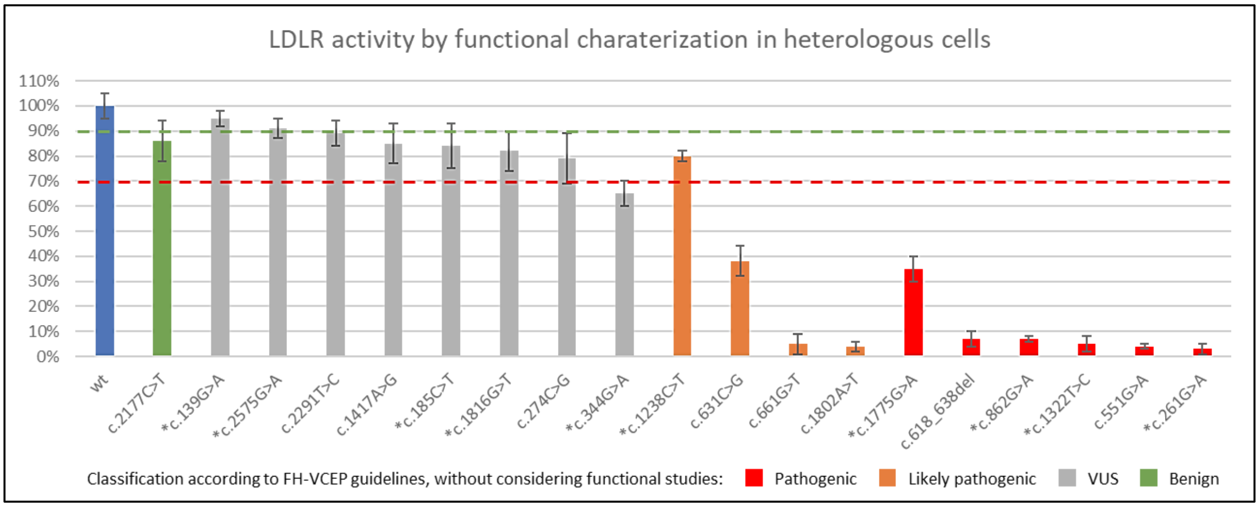


**Supplementary Figure 1**. Functional characterization of LDLR activity in heterologous cells transfected with mutant plasmids. LDLR activity levels adapted from references[2–6]. The lowest activity value among LDLR expression, LDL binding and LDL internalization is displayed. The green dotted line represents the recommended LDLR activity threshold (>90% compared to wild-type) for application of Level 1 BS3 functional study evidence, and the red dotted line represents the recommended LDLR activity threshold (<70% compared to wild-type) for application of Level 1 PS3 functional study evidence. Note: the star (*) represents variants included our pilot study, where the associated classification does not require the application of PS3/BS3 criteria. For the remaining variants, classifications were performed by only one curator and using internal case-level data, so final FH VCEP classifications which appear in ClinVar may be different.

**Supplementary Table 4**. *LDLR* cysteine residues involved in disulfide bond formation.

| Residue | Domain | Structure analysis | Predicted impact on LDLR structure and/or function |
|---|---|---|---|
| p.Cys27 | LDL-receptor class A 1 | disulfide bond | folding defect |
| p.Cys34 | LDL-receptor class A 1 | disulfide bond | folding defect |
| p.Cys39 | LDL-receptor class A 1 | disulfide bond | folding defect |
| p.Cys46 | LDL-receptor class A 1 | disulfide bond | folding defect |
| p.Cys52 | LDL-receptor class A 1 | disulfide bond | folding defect |
| p.Cys63 | LDL-receptor class A 1 | disulfide bond | folding defect |
| p.Cys68 | LDL-receptor class A 2 | disulfide bond | folding defect |
| p.Cys75 | LDL-receptor class A 2 | disulfide bond | folding defect |
| p.Cys82 | LDL-receptor class A 2 | disulfide bond | folding defect |
| p.Cys89 | LDL-receptor class A 2 | disulfide bond | folding defect |
| p.Cys95 | LDL-receptor class A 2 | disulfide bond | folding defect |
| p.Cys104 | LDL-receptor class A 2 | disulfide bond | folding defect |
| p.Cys109 | LDL-receptor class A 3 | disulfide bond | folding defect; LDL binding defect |
| p.Cys116 | LDL-receptor class A 3 | disulfide bond | folding defect; LDL binding defect |
| p.Cys121 | LDL-receptor class A 3 | disulfide bond | folding defect; LDL binding defect |
| p.Cys128 | LDL-receptor class A 3 | disulfide bond | folding defect; LDL binding defect |
| p.Cys134 | LDL-receptor class A 3 | disulfide bond | folding defect; LDL binding defect |
| p.Cys143 | LDL-receptor class A 3 | disulfide bond | folding defect; LDL binding defect |
| p.Cys148 | LDL-receptor class A 4 | disulfide bond; acidic pH intramolecular binding interface | folding defect; receptor-recycling defect; LDL binding defect |
| p.Cys155 | LDL-receptor class A 4 | disulfide bond | folding defect; LDL binding defect |
| p.Cys160 | LDL-receptor class A 4 | disulfide bond; acidic pH intramolecular binding interface | folding defect; receptor-recycling defect; LDL binding defect |
| p.Cys167 | LDL-receptor class A 4 | disulfide bond | folding defect; LDL binding defect |
| p.Cys173 | LDL-receptor class A 4 | disulfide bond; acidic pH intramolecular binding interface | folding defect; receptor-recycling defect; LDL binding defect |
| p.Cys184 | LDL-receptor class A 4 | disulfide bond | folding defect; LDL binding defect |
| p.Cys197 | LDL-receptor class A 5 | disulfide bond | folding defect; LDL binding defect |
| p.Cys204 | LDL-receptor class A 5 | disulfide bond | folding defect; LDL binding defect |
| p.Cys209 | LDL-receptor class A 5 | disulfide bond | folding defect; LDL binding defect |
| p.Cys216 | LDL-receptor class A 5 | disulfide bond | folding defect; LDL binding defect |
| p.Cys222 | LDL-receptor class A 5 | disulfide bond; acidic pH intramolecular binding interface | folding defect; receptor-recycling defect; LDL binding defect |
| p.Cys231 | LDL-receptor class A 5 | disulfide bond | folding defect; LDL binding defect |
| p.Cys236 | LDL-receptor class A 6 | disulfide bond | folding defect; LDL binding defect |
| p.Cys243 | LDL-receptor class A 6 | disulfide bond | folding defect; LDL binding defect |
| p.Cys248 | LDL-receptor class A 6 | disulfide bond | folding defect; LDL binding defect |
| p.Cys255 | LDL-receptor class A 6 | disulfide bond | folding defect; LDL binding defect |

| | | | |
|---|---|---|---|
| p.Cys261 | LDL-receptor class A 6 | disulfide bond | folding defect; LDL binding defect |
| p.Cys270 | LDL-receptor class A 6 | disulfide bond | folding defect; LDL binding defect |
| p.Cys276 | LDL-receptor class A 7 | disulfide bond | folding defect; LDL binding defect |
| p.Cys284 | LDL-receptor class A 7 | disulfide bond | folding defect; LDL binding defect |
| p.Cys289 | LDL-receptor class A 7 | disulfide bond | folding defect; LDL binding defect |
| p.Cys296 | LDL-receptor class A 7 | disulfide bond | folding defect; LDL binding defect |
| p.Cys302 | LDL-receptor class A 7 | disulfide bond | folding defect; LDL binding defect |
| p.Cys313 | LDL-receptor class A 7 | disulfide bond | folding defect; LDL binding defect |
| p.Cys318 | EGF-like 1 | disulfide bond | folding defect; LDL binding defect |
| p.Cys325 | EGF-like 1 | disulfide bond | folding defect; LDL binding defect |
| p.Cys329 | EGF-like 1 | disulfide bond | folding defect; LDL binding defect |
| p.Cys338 | EGF-like 1 | disulfide bond | folding defect; LDL binding defect |
| p.Cys340 | EGF-like 1 | disulfide bond | folding defect; LDL binding defect |
| p.Cys352 | EGF-like 1 | disulfide bond | folding defect; LDL binding defect |
| p.Cys358 | EGF-like 2; calcium-binding | disulfide bond | folding defect; receptor-recycling defect |
| p.Cys364 | EGF-like 2; calcium-binding | disulfide bond | folding defect; receptor-recycling defect |
| p.Cys368 | EGF-like 2; calcium-binding | disulfide bond | folding defect; receptor-recycling defect |
| p.Cys377 | EGF-like 2; calcium-binding | disulfide bond | folding defect; receptor-recycling defect |
| p.Cys379 | EGF-like 2; calcium-binding | disulfide bond | folding defect; receptor-recycling defect |
| p.Cys392 | EGF-like 2; calcium-binding | disulfide bond | folding defect; receptor-recycling defect |
| p.Cys667 | EGF-like 3 | disulfide bond | folding defect; receptor-recycling defect |
| p.Cys677 | EGF-like 3 | disulfide bond | folding defect; receptor-recycling defect |
| p.Cys681 | EGF-like 3 | disulfide bond | folding defect; receptor-recycling defect |
| p.Cys696 | EGF-like 3 | disulfide bond | folding defect; receptor-recycling defect |
| p.Cys698 | EGF-like 3 | disulfide bond | folding defect; receptor-recycling defect |
| p.Cys711 | EGF-like 3 | disulfide bond | folding defect; receptor-recycling defect |

Adapted from Guo et al., 2019.[7] Residues correspond to *LDLR* transcript NM_000527.5.
Abbreviations: Cys, cysteine; EGF, epidermal growth factor

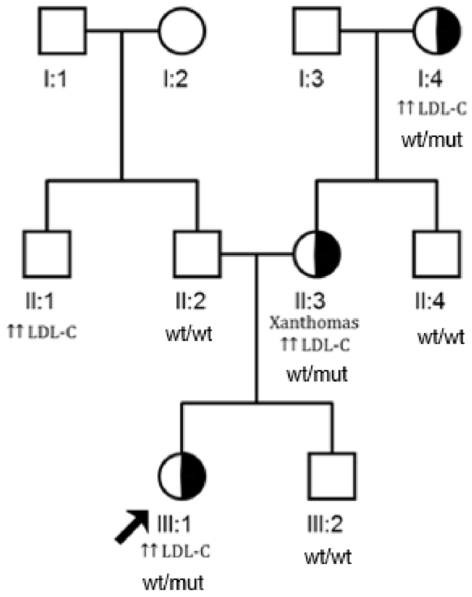


**Supplementary Figure 2.** Pedigree of a FH family. Index case is identified with an arrow. Half-filled symbols represent heterozygous individuals. Index case III:1 inherited her *LDLR* variant from the maternal (II:3) side of the family. Her father (II:2) has normal cholesterol, no cardiovascular disease history, and is negative for the *LDLR* variant; therefore, her father (II:2) and paternal uncle (II:1) should not be considered in the co-segregation study. Similarly, the maternal grandfather (I:3) should not be considered. In this family the individuals that can be considered informative meioses are the index case's brother (III:2), mother (II:3), maternal uncle (II:4) and maternal grandmother (I:4). Index cases should not be counted as positive cases for co-segregation results.

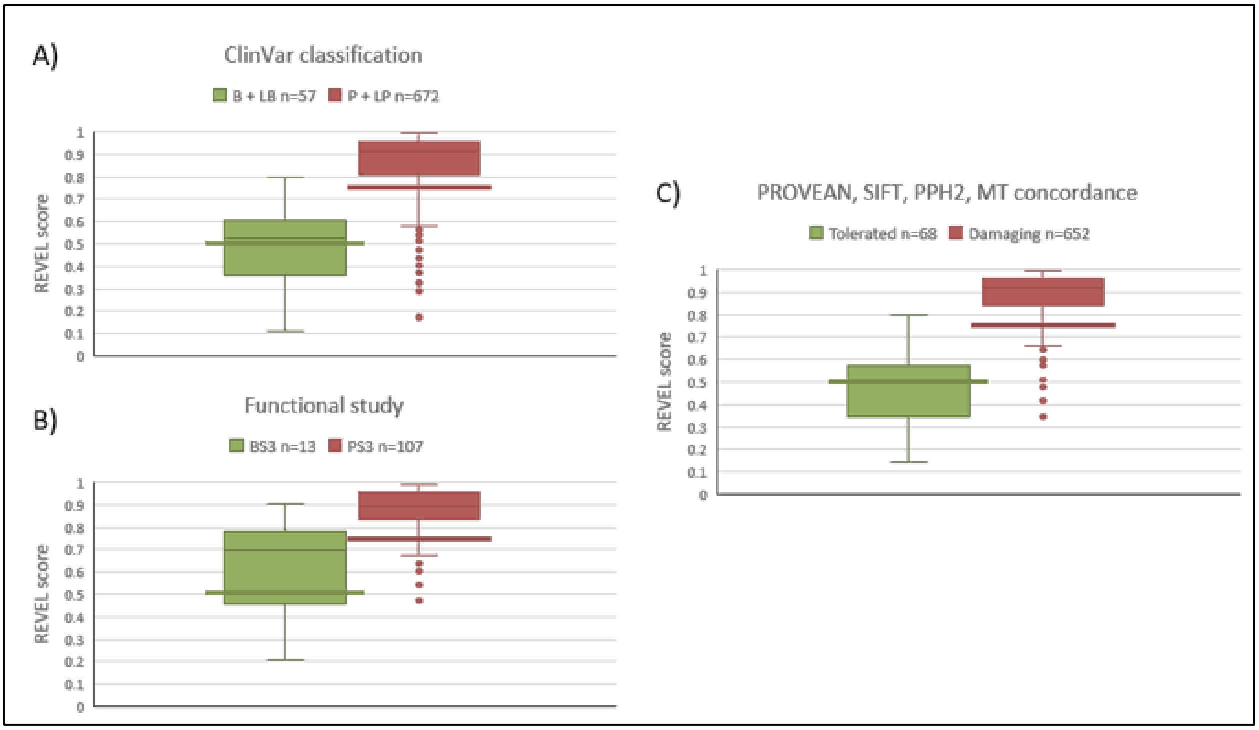


**Supplementary Figure 3**. REVEL score distributions for determining missense PP3 and BP4 thresholds in *LDLR*. **A)** REVEL scores for *LDLR* missense variants classified as Benign/Likely benign (B+LB) or Pathogenic/Likely pathogenic (P+LP) in ClinVar. **B)** REVEL scores for *LDLR* missense variants with neutral or damaging BS3/PS3 evidence from functional studies. **C)** REVEL scores for *LDLR* missense variants with concordant *in silico* results for Poly-Phen-2 (PPH2), SIFT, PROVEAN and MutationTaster (MT). The green line represents the suggested threshold score of <0.50 used in the applicability of ACMG/AMP criterion BP4, and the red line represents the suggested threshold score of >0.75 used in the applicability of ACMG/AMP criterion PP3.

**Supplementary Table 5**. *LDLR* pilot study variants.

| ***LDLR* Variant (NM_000527.5)** | **ClinVar ID** | **Previous ClinVar Status*** | **ClinVar Classification** | **Pilot Study Classification** |
|---|---|---|---|---|
| c.1061-?_1845+?del | 265901 | 1-star | P | Pathogenic |
| c.1A>T (p.Met1Leu) | 250968 | 1-star | LP | VUS |
| c.58G>A (p.Gly20Arg) | 161272 | 1-star | Conflicting: B (1); LB (5); VUS (5) | Benign |
| c.139G>A (p.Asp47Asn) | 251034 | 1-star | Conflicting: LP (1); VUS (2) | VUS |
| c.185C>G (p.Thr62Arg) | 375775 | 1-star | LP | VUS |
| c.185C>T (p.Thr62Met) | 161273 | 1-star | Conflicting: LB (1); VUS (7) | VUS |
| c.232C>T (p.Arg78Cys) | 161289 | 1-star | Conflicting: LP (1); VUS (1) | VUS |
| c.259T>G (p.Trp87Gly) | 3685 | 2-star | P/LP | Pathogenic |
| c.261G>A (p.Trp87Ter) | 251100 | 2-star | P | Pathogenic |
| c.268G>T (p.Asp90Tyr) | 251106 | 2-star | LP | Likely pathogenic |
| c.296C>G (p.Ser99Ter) | 161269 | 2-star | P | Pathogenic |
| c.313+1G>A | 3736 | 1-star | Conflicting: LB (1); LP (2); P (12) | Pathogenic |
| c.343C>T (p.Arg115Cys) | 251162 | 1-star | Conflicting: LP (1); P (1); VUS (1) | Likely pathogenic |
| c.344G>A (p.Arg115His) | 225402 | 1-star | Conflicting: LB (1); LP (2); P (1); VUS (3) | VUS |
| c.693C>G (p.Cys231Trp) | 251400 | 2-star | LP | Likely pathogenic |
| c.718G>T (p.Glu240Ter) | 251422 | 2-star | P | Pathogenic |
| c.757C>T (p.Arg253Trp) | 161261 | 1-star | Conflicting: LB (2); LP (1); P (1); VUS (4) | VUS |
| c.798T>A (p.Asp266Glu) | 161287 | 1-star | Conflicting: LB (1); LP (9); P (6); VUS (2) | Pathogenic |
| c.806G>A (p.Gly269Asp) | 161279 | 1-star | Conflicting: B (1); LB (8); LP (1); VUS (1) | Likely benign |
| c.862G>A (p.Glu288Lys) | 161268 | 1-star | Conflicting: LP (9); P (1); VUS (1) | Pathogenic |
| c.907C>T (p.Arg303Trp) | 161281 | 1-star | Conflicting: LP (1); VUS (4) | VUS |
| c.910G>T (p.Asp304Tyr) | 251517 | 2-star | P/LP | Likely pathogenic |
| c.967G>A (p.Gly323Ser) | 161282 | 1-star | Conflicting: LP (2); VUS (1) | VUS |
| c.970G>A (p.Gly324Ser) | 161263 | 1-star | Conflicting: B (4); LB (3); LP (1); P (1); VUS (2) | Benign |
| c.1024G>T (p.Asp342Tyr) | 251603 | 1-star | Conflicting: LP (1); VUS (3) | VUS |
| c.1055G>A (p.Cys352Tyr) | 36450 | 2-star | P/LP | Likely pathogenic |
| c.1060+10G>C | 226709 | 2-star | B/LB | Benign |
| c.1171G>A (p.Ala391Thr) | 183138 | 2-star | B/LB | Benign |

| | | | | |
|---|---|---|---|---|
| c.1186+5G>A | 251706 | 1-star | Conflicting: LP (2); P (1); VUS (2) | Likely pathogenic |
| c.1216C>A (p.Arg406=) | 3746 | 2-star | P | Likely pathogenic |
| c.1217G>A (p.Arg406Gln) | 228798 | 1-star | Conflicting: LP (1); P (1); VUS (3) | Likely pathogenic |
| c.1222G>A (p.Glu408Lys) | 36453 | 2-star | P/LP | Likely pathogenic |
| c.1238C>T (p.Thr413Met) | 161276 | 1-star | Conflicting: LP (6); VUS (2) | Likely pathogenic |
| c.1322T>C (p.Ile441Thr) | 251783 | 2-star | P/LP | Pathogenic |
| c.1323C>T (p.Ile441=) | 456650 | 2-star | B/LB | Benign |
| c.1576C>T (p.Pro526Ser) | 183120 | 1-star | Conflicting: LP (4); P (2); VUS (1) | VUS |
| c.1775G>A (p.Gly592Glu) | 161271 | 2-star | P/LP | Pathogenic |
| c.1783C>T (p.Arg595Trp) | 161290 | 1-star | Conflicting: LP (5); P (3); VUS (3) | Pathogenic |
| c.1816G>T (p.Ala606Ser) | 161264 | 2-star | VUS | VUS |
| c.1855T>C (p.Phe619Leu) | 252083 | 1-star | Conflicting: LP (1); VUS (3) | Likely pathogenic |
| c.1965C>G (p.Phe655Leu) | 252135 | 1-star | Conflicting: LB (1); LP (1); P (1) | Likely pathogenic |
| c.1966C>A (p.His656Asn) | 252136 | 1-star | Conflicting: B (2); LP (2) | VUS |
| c.2000G>A (p.Cys667Tyr) | 3689 | 2-star | P/LP | Likely pathogenic |
| c.2043C>G (p.Cys681Trp) | 252188 | 1-star | LP | Pathogenic |
| c.2096C>T (p.Pro699Leu) | 252219 | 1-star | Conflicting: LP (7); P (2); VUS (2) | VUS |
| c.2101G>A (p.Gly701Ser) | 183130 | 1-star | Conflicting: B (1); LB (2); LP (1); VUS (3) | VUS |
| c.2140+1G>A | 3744 | 2-star | P/LP | Pathogenic |
| c.2140+5G>A | 36460 | 1-star | Conflicting: B (7); LB (6); VUS (1) | Benign |
| c.2389+4A>G | 252304 | 1-star | Conflicting: LB (1); VUS (3) | VUS |
| c.2389+8C>T | 413774 | 2-star | B/LB | VUS |
| c.2479G>A (p.Val827Ile) | 36462 | 1-star | Conflicting: B (3); LB (3); LP (1); VUS (7) | VUS |
| c.2531G>A (p.Gly844Asp) | 3734 | 1-star | LP | Likely pathogenic |
| c.2546C>A (p.Ser849Ter) | 252350 | 2-star | P/LP | Likely pathogenic |
| c.2575G>A (p.Val859Met) | 252360 | 1-star | Conflicting: LB (2); VUS (2) | Likely benign |

Abbreviations: B, benign; LB, likely benign; VUS, variant of uncertain significance; LP, likely pathogenic; P. pathogenic. *ClinVar status prior submitting FH VCEP classifications using the approved *LDLR*-specific ACMG/AMP guidelines; these variants are now at 3-star status.

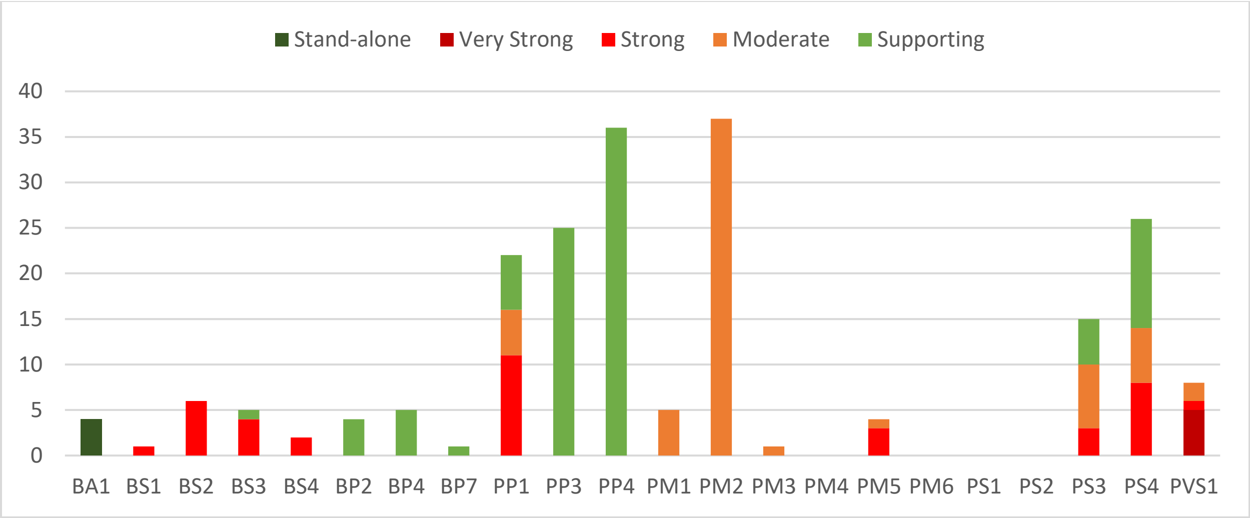


**Supplementary Figure 4**. Number of times each ACMG/AMP criteria code was applied in a pilot study of 54 *LDLR* variants.

### Supplementary File References
